# Which social media platforms facilitate monitoring the opioid crisis?

**DOI:** 10.1101/2024.07.06.24310035

**Authors:** Kristy A. Carpenter, Anna T. Nguyen, Delaney A. Smith, Issah A. Samori, Keith Humphreys, Anna Lembke, Mathew V. Kiang, Johannes C. Eichstaedt, Russ B. Altman

## Abstract

Social media can provide real-time insight into trends in substance use, addiction, and recovery. Prior studies have used platforms such as Reddit and X (formerly Twitter), but evolving policies around data access have threatened these platforms’ usability in research. We evaluate the potential of a broad set of platforms to detect emerging trends in the opioid epidemic. From these, we created a shortlist of 11 platforms, for which we documented official policies regulating drug-related discussion, data accessibility, geolocatability, and prior use in opioid-related studies. We quantified their volumes of opioid discussion, capturing informal language by including slang generated using a large language model. Beyond the most commonly used Reddit and X, the platforms with high potential for use in opioid-related surveillance are TikTok, YouTube, and Facebook. Leveraging many different social platforms, instead of a single platform, safeguards against sudden changes to data access and may better capture all populations that use opioids than any single platform.

**Teaser:** TikTok, Facebook, and YouTube may complement Reddit and X as text sources to monitor trends in the opioid epidemic.

## Introduction

Real-time tracking of substance use trends is key to understanding epidemics of addiction, including the North American opioid crisis. Heterogeneity in prominent substances, drug availability, and usage patterns across the epidemic require different intervention strategies, and policy makers must be sensitive to these dynamics. Academic and government surveys are standard practice for tracking usage patterns. However, difficulties with self-reported opioid use limit the reliability of these surveys, leading to inaccurate estimates of the prevalence and nature of opioid use (*1*). Moreover, surveys can take months or years to complete and results may not reflect quickly evolving trends in opioid use.

Social media platforms are an alternative data source which might be used to track patterns of opioid use. Unlike official overdose statistics, social media provides real-time, high-volume, and widely accessible streams of information (*2*, *3*). These platforms capture unfiltered experiences across diverse populations, including encounters with substances that might not otherwise be disclosed (*4*). Thus, social media can aid in identification and geolocation of emerging drugs and in tracking practices among people who use opioids (PWUOs). Understanding these patterns could help policymakers anticipate hotspots for overdoses.

Previous work has used social media data at the individual level to detect opioid misuse (*4*), flag indicators of addiction (*5*), and assess individual risk of relapse (*6*). A larger body of work is dedicated to tracking population trends in opioid misuse (*7*) and opioid-related mortality (*8*, *9*). Language use on these platforms may be more predictive of trends in county-level deaths than factors such as demographics, healthcare access, and physical pain (*8*). Other work has triangulated social media data with other surveillance datasets (*e.g.* emergency department admissions) to develop holistic models (*10*).

Some prior research found correlations between opioid mentions on social media and opioid use and overdose without distinguishing direct disclosures of opioid use from mentions referring to the opioid epidemic generally, popular culture, or the drug use of others (*2*, *8–10*). Relying only upon explicit disclosures of a social media user’s illicit opioid use may not reflect true rates of opioid usage as many PWUO will not post about their experiences, and some social media users may post false stories of drug use. Additionally, discussion of opioids in the broader community can yield insight into general themes and sentiments regarding the opioid epidemic, such as stigma or the reception of opioid policy.

Most prior work uses only a few social media platforms, chiefly X (formerly Twitter) and Reddit. However, access to these data sources is dependent on corporate decisions. For example, Pushshift (*12*) was a popular research tool for extracting Reddit posts, but Reddit started to limit API requests for third-party data access in 2023, effectively disabling Pushshift. A similar phenomenon occurred with the X API contemporaneously. As digital platforms evolve, it is critical to understand the scope of available datasets and potential alternatives. Additionally, different social media platforms have different constituent demographics; no individual platform fully represents the population at large. An effort combining multiple platforms may be better able to capture trends in this crisis.

There are no comprehensive studies evaluating the nature, volume, and quality of opioid-related discussions across social media platforms. Although there are several systematic literature reviews of this field (*3*, *13*), a direct evaluation of a broad range of social media platforms is needed to best leverage available social media platforms.

Although opioids contribute to morbidity and mortality in many countries, the United States and Canada have an unusually serious epidemic of addiction and overdose, which was triggered by opioid overprescription and is now mainly driven by fentanyl use (*14–19*). Therefore, while social media promises to advance understanding of opioid use worldwide, it is particularly urgent to leverage to address the North American opioid epidemic.

Here, we identified social media platforms that may be suitable for text-based opioid research and characterized their opioid-related discussions. We created a shortlist of eleven platforms for which we investigated censorship policies, data accessibility, and prior use in opioid research. We discuss the utility, availability, and stability of these platforms for the purpose of informing design of a social media early-warning system for trends in the North American opioid epidemic. We use all opioid mentions to capture the full potential of a platform to yield insight on different aspects of opioid research. While this work is limited to the United States and Canada, and the English language, we provide a framework to conduct similar work in other regions and languages.

## Results

### Identifying social media platforms

We took the union of the platforms found from the sources listed in Methods to create a superset of 71 candidate platforms (**Supplementary Data S1**).

### Creating platform shortlist

We applied our shortlisting criteria to the 71 platforms iteratively (**Figure 1**, **Table S2**). Four platforms were not active as of July 2023. Twenty-one active platforms did not meet our definition of social media and 11 active platforms were private messaging platforms. Eleven active social media sites were not based in the United States or did not have English as the default language for a user in the United States. Among the remaining sites, 13 platforms returned fewer than 25,000 query results on the selected opioid keywords.

**Fig. 1.**
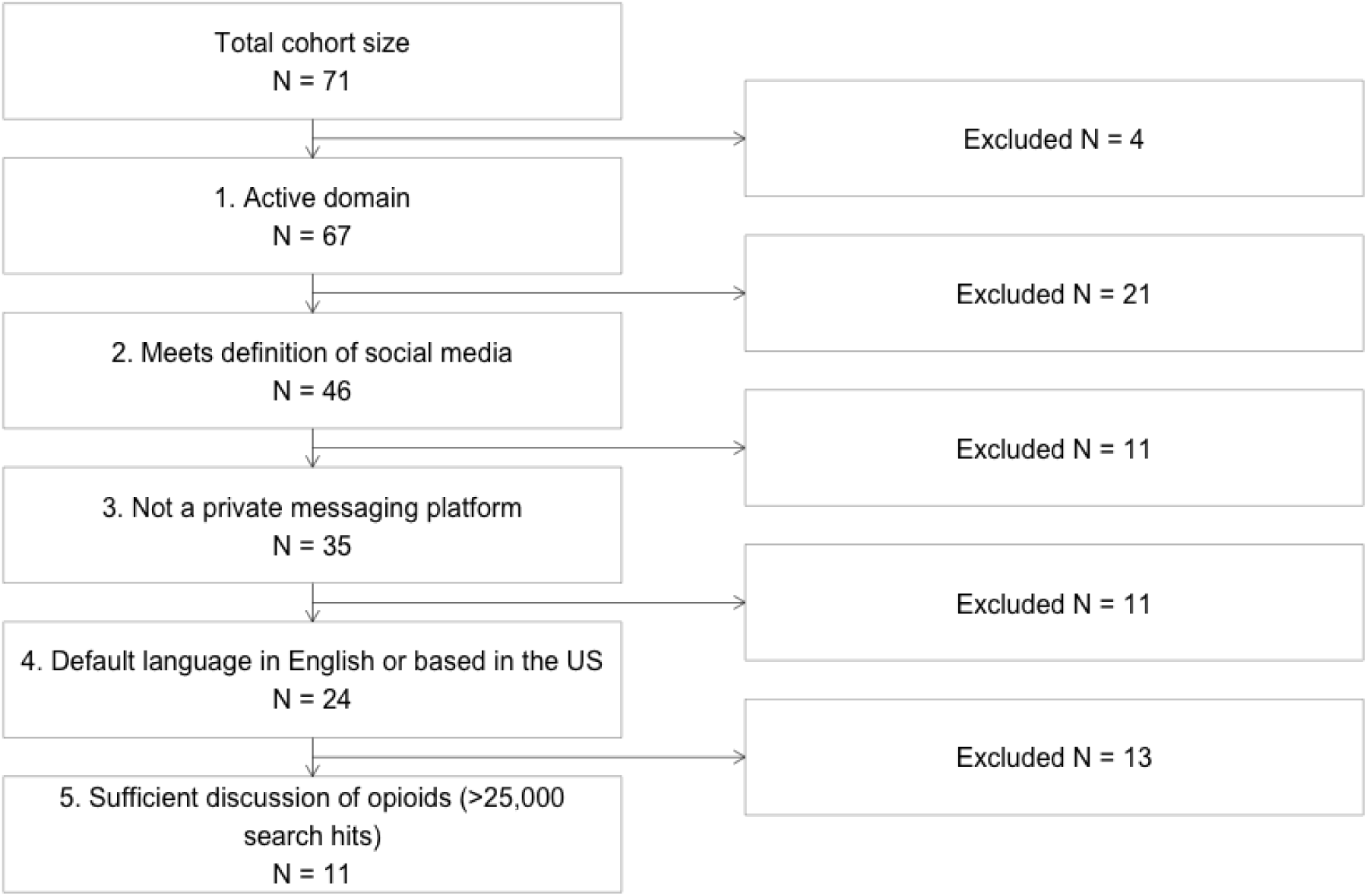
Platform shortlisting consort diagram. Consort diagram of iterative exclusion criteria to attain shortlist of 11 social media platforms for further characterization.

This process left 11 (15.5%) platforms for further evaluation: Bluelight, drugs-forum.com, Facebook, Instagram, LinkedIn, Pinterest, Reddit, TikTok, Tumblr, X (formerly Twitter), and YouTube.

Characteristics of these platforms are described in **Table 1**.

**Table 1.**
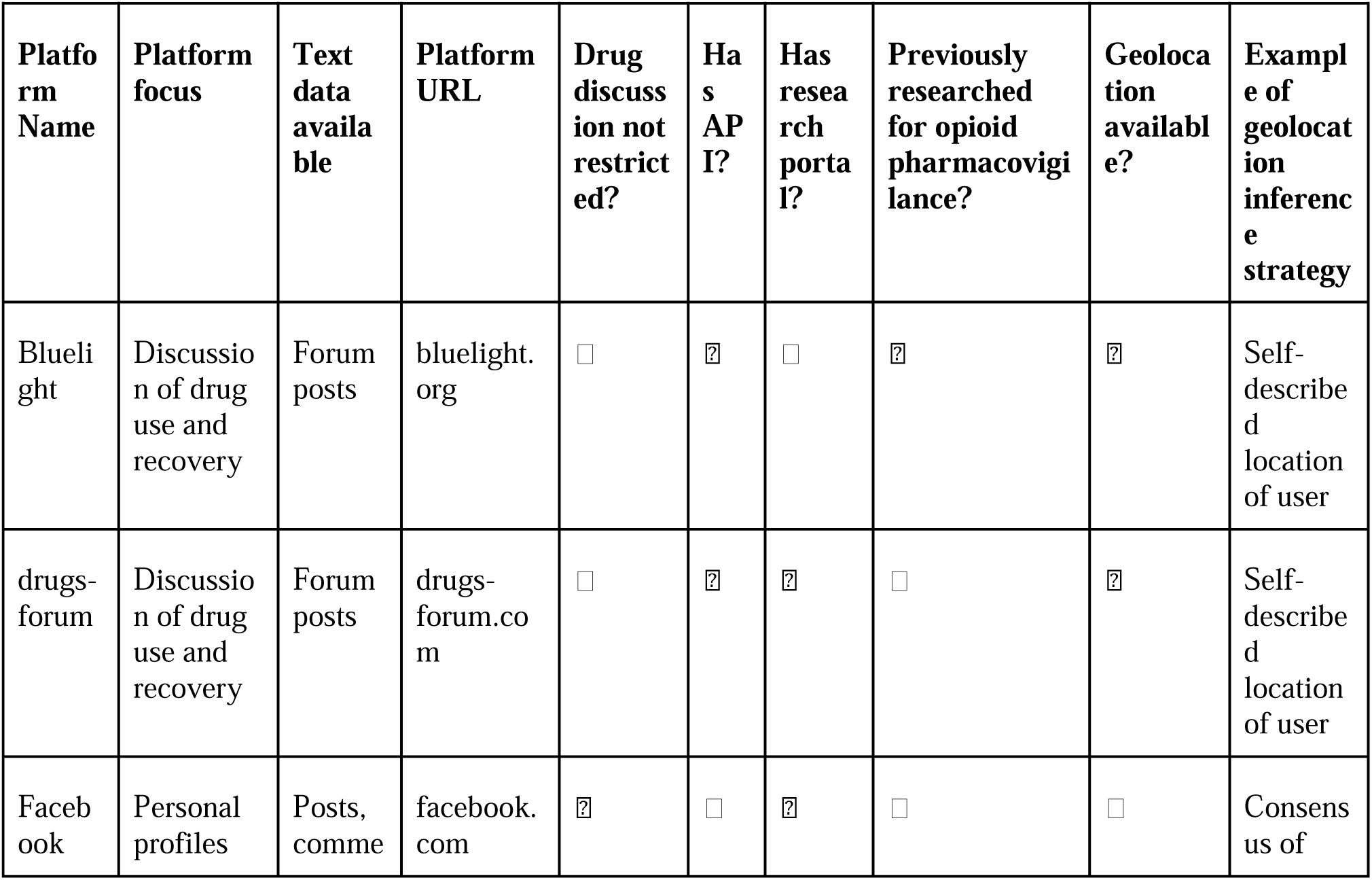
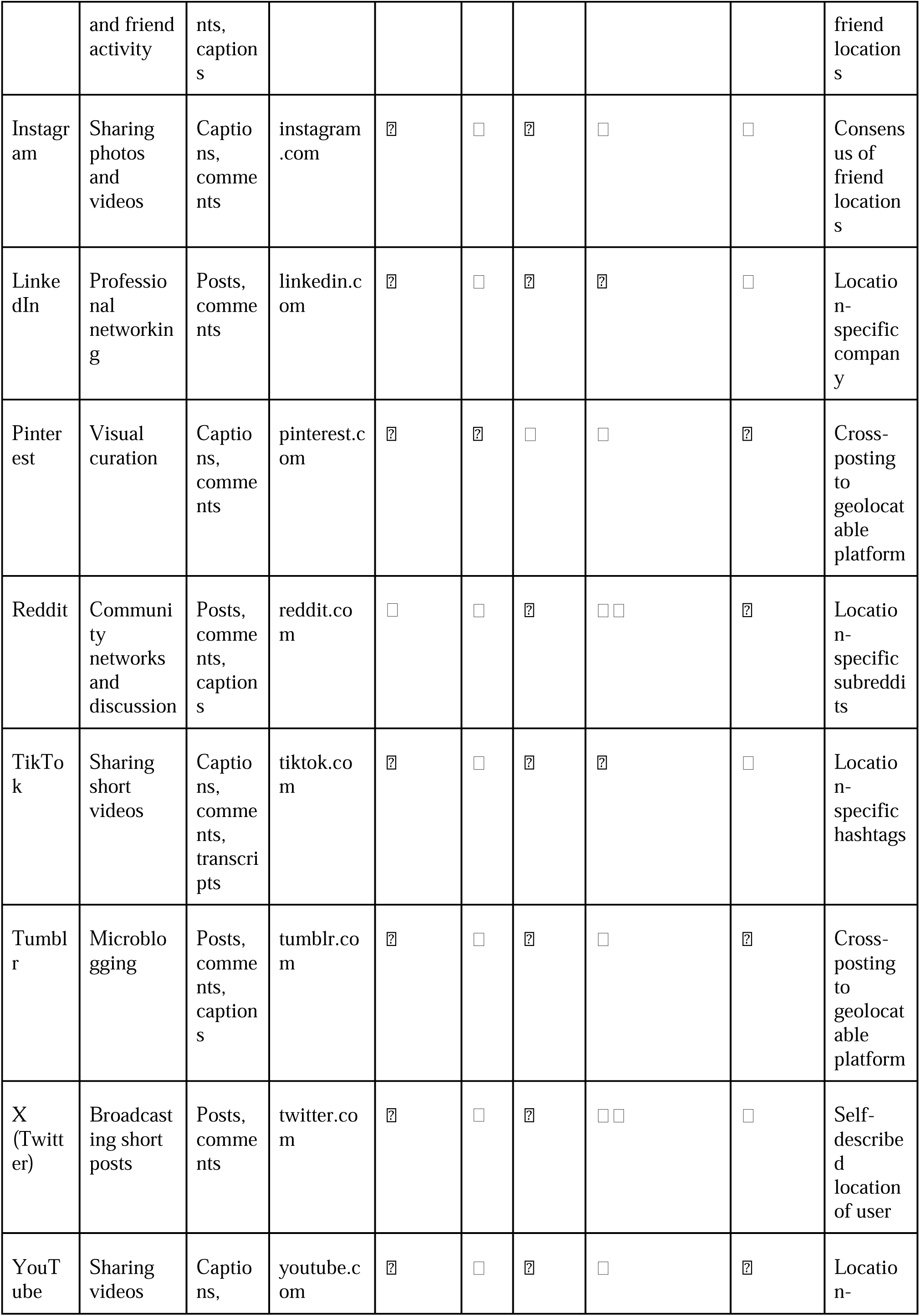
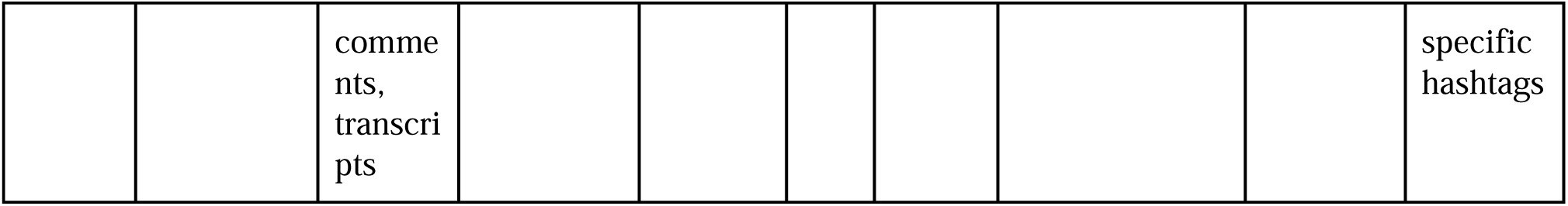
Summary of characteristics of shortlisted social media platforms. “Platform focus” gives a brief description of the primary usage of the platform. “Text data available” lists the types of text content that the platform contains, as determined by manual inspection of platforms. “Drug discussion not restricted” column has a checkmark if the platform does not restrict drug-related discussion, and an X if the platform has some form of restrictions, as determined by inspection of platform terms of use (provided in **Supplementary Text S7**). “Has API” column has a checkmark if the platform has an API available (whether the API is freely available or requires authorization before access), and an X if not. “Has research portal” column has a checkmark if the platform has a non-API platform for acquisition of platform data or to receive more information about collaboration with the platform, and an X if not. API and research portal designations were determined by inspection of platform data availability (provided in **Supplementary Text S8**). “Previously researched for opioid pharmacovigilance” column reflects the relative amount of prior research related to opioid surveillance, with a checkmark indicating some prior use in the literature, two checkmarks indicating high prior use in the literature, and an X indicating no prior use in the literature (see “Prior Use in Research Literature” section of **Results**). “Geolocation available” column has a checkmark if explicit geolocation data is provided for *any* platform content (this does not indicate explicit geolocation available for all content), and an X if not, as determined by inspection of public-facing platform data specifications (see “Evaluating data accessibility for academic research purposes” section of **Results**). “Example of geolocation inference strategy” column provides one possible method for inferring geolocation of platform content, based on inference strategies previously employed in the broader literature (see “Evaluating data accessibility for academic research purposes section” of **Results**).

### Measuring the volume of opioid-related discussion

The type and volume of publicly available opioid-related text varied across platforms (Figure 2, Figure 3, Figure S1, Table S3, Table S4). We observed the highest total volume of opioid-related discussion on YouTube and Facebook (Figure 2a, Figure S1, Table S3). These were followed by LinkedIn, TikTok, and Reddit. The platforms with the lowest total volume of opioid-related discussion were Bluelight and drugs-forum.

**Fig. 2.**
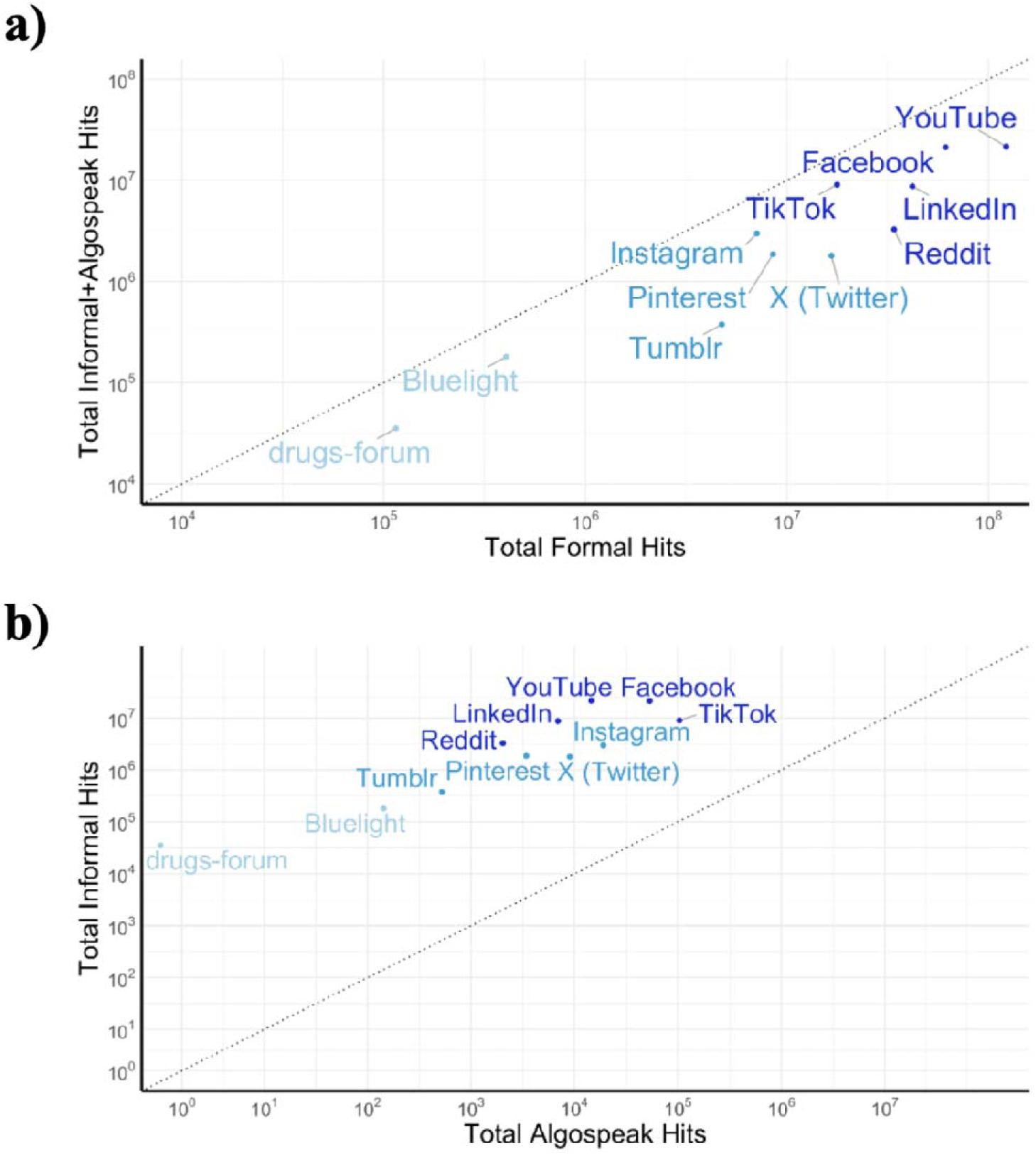
Total term hits for all shortlisted platforms. Shown with a logarithmic scale. Platforms are clustered into three categories by volume of hits, shown by color (darker shade of blue indicates higher volume of hits). Dotted line shows y=x diagonal. **(A)** Total hits for formal opioid term list versus the sum of the total hits for the informal opioid term list and the algospeak opioid term list. **(B)** Total hits for informal opioid term list versus algospeak opioid term list.

**Fig. 3.**
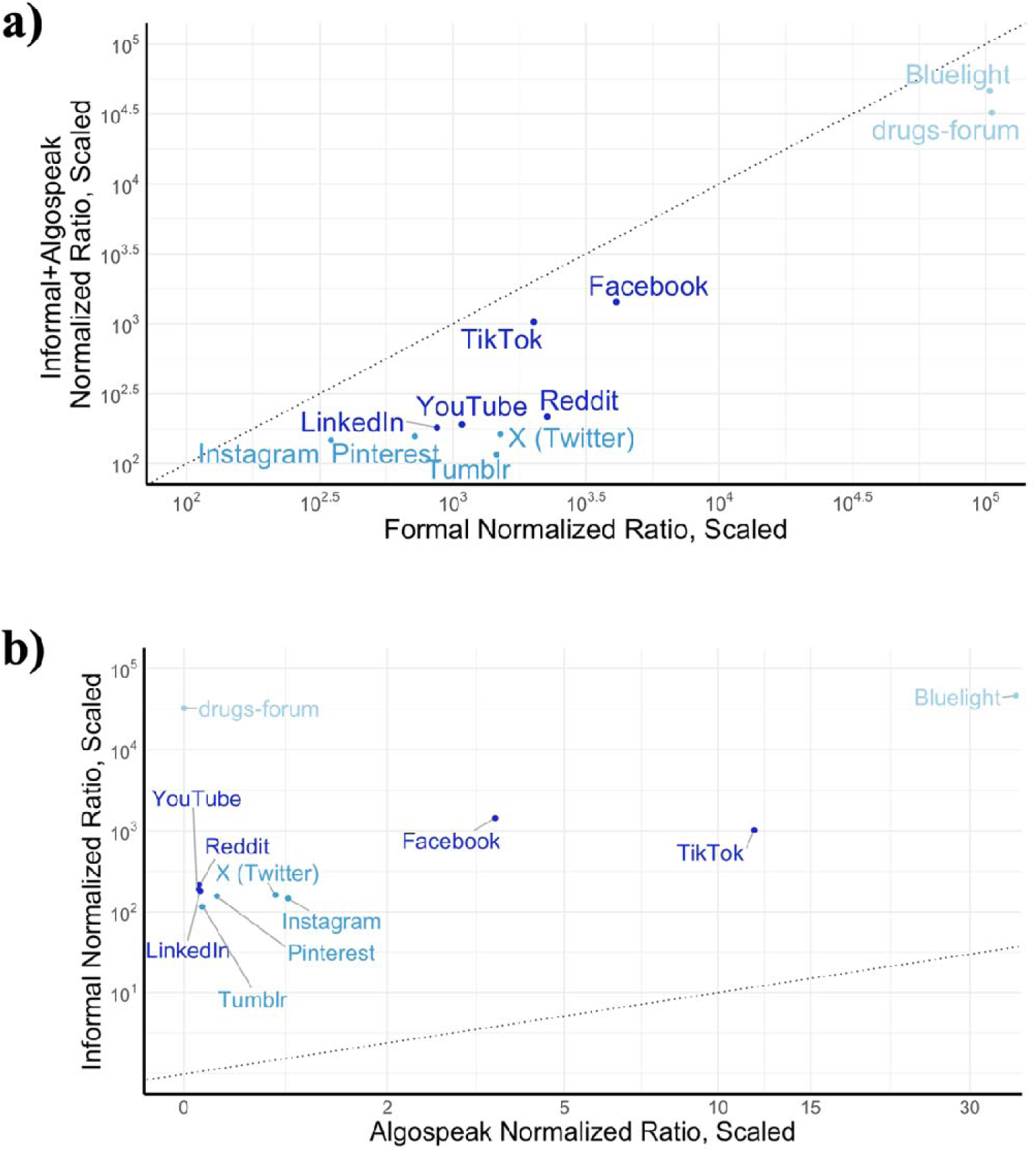
Term hits normalized by number of hits for common nouns. Shown with a logarithmic scale and scaling factor of 100,000. As in Figure 2, platforms are clustered into three categories by total volume of hits, shown by color (darker shade of blue indicates higher volume of hits). Dotted line shows y=x diagonal. **(A)** Normalized hit ratios for formal opioid term list versus for informal opioid term list and algospeak opioid term list. **(B)** Normalized hit ratios for informal opioid term list versus algospeak opioid term list.

Most platforms followed a linear relationship between the amount of formal opioid terms and the amount of informal (including algospeak) opioid terms. TikTok and Instagram skewed toward more informal term hits, and LinkedIn, Reddit, and X skewed toward more formal term hits.

The relationship between the total number of informal hits versus the total number of algospeak hits revealed similar trends (Figure 2b). The outliers reveal which platforms have more algospeak, indicating a response to censorship. TikTok by far had the most amount of algospeak. Instagram and X also skewed toward algospeak. LinkedIn and Reddit both skewed away from algospeak.

While having less drug-related discussion overall, Bluelight and drugs-forum had dramatically higher rates of drug discussion relative to non-drug discussion (**Figure 3a**). Among the social media platforms with general scope, TikTok and Facebook had the highest relative amount of drug discussion compared to non-drug discussion.

We visualized the relative amounts of informal and algospeak term hits for the 11 platforms (**Figure 3b**). Bluelight and drugs-forum show large separation from the nine general social media platforms, especially with respect to informal (non-algospeak) terms. Of the nine general social media platforms, Facebook and TikTok had the highest relative amounts of drug discussion with respect to both informal and algospeak term hits.

### Content restrictions and censorship policies

All eleven shortlisted platforms state expectations regarding drug-related discussion (**Supplementary Text S7**). High-level classifications of content policies are shown in **Table 1**.

All eleven platforms state that the sale of illicit substances is not allowed. Facebook, Instagram, TikTok, and YouTube state that posting content related to recreational use of drugs is prohibited. TikTok underscores that such content is dangerous for young people. These platforms make exceptions for recovery-oriented or educational content. LinkedIn prohibits depictions of “drug abuse.” Reddit includes communities focused on drug-related discussion, which are age-restricted. Bluelight and drugs-forum are distinct as forums dedicated to drug-related discussion. Both platforms state that they are safe spaces for discussion of all aspects of drug use and recovery.

### Evaluating data accessibility for academic research purposes

Many platforms provide Application Programming Interfaces (APIs) for data extraction. Generally, APIs are available in tiers where more expensive versions provide more comprehensive access. Research APIs often provide free or discounted access to users affiliated with a research institute. In some cases, outside groups have maintained third-party APIs (*e.g.* Pushshift (*12*) for Reddit). We found that smaller drug-focused forums do not have APIs that allow for easy data extraction. We provide an overview of data access capabilities in **Table 1** and per-platform details in **Supplementary Text S8**.

An early-warning system for trends in the opioid epidemic requires geolocatability. The geolocation of social media content can be obtained directly from the platform or, if unavailable, inferred. Some social media platforms provide geolocation of users or posts. X (formerly Twitter) gives users the option to geotag their tweets. A small subset of users opt to do this (under 2% (*20*)). Many groups have leveraged geotagged tweets for opioid-related research (*21–27*) and other research areas (*28*, *29*). It was possible to geotag a tweet with latitude and longitude coordinates until June 2019; since then, users can only tag tweets with place objects that have coordinate bounding boxes (*20*, *30*, *31*).

Similarly, Facebook (*32*), Instagram (*33–36*), and YouTube (*37–39*) give users the option to tag their content with geolocations. However, these tags can be used for purposes other than reporting the location from where the post was made (*40*). Facebook has a Data for Good program (*41*) which includes province-level GPS data from a subset of consenting users; previous work leveraged this data to study phenomena related to the COVID-19 pandemic (*42*, *43*). State *et al.* used provided user and company locations to track migration patterns using LinkedIn (*44*).

The geolocatability of TikTok users has been prominently discussed in the US (*45–47*). Like other social media platforms, TikTok uses user location to personalize their content feed and allows users to add location tags to their videos (*48*). Various groups have characterized the privacy aspect of geolocation on TikTok (*49*, *50*), but there is little research using tagged locations for geographically-informed analysis. Zanettou *et al.* recruited consenting users to donate their geotagged data, which each user can request under the EU’s GDPR regulation (*51*).

Many online forums allow users to indicate their location on their profile. This is required on drugs-forum and optional on Bluelight. These location entries are unstandardized free text and can be anything from cities (*e.g.* “palo alto”, “Palo Alto, CA”), states (*e.g.* “CA, USA”, “California”, “cali”), regions (*e.g.* “NorCal”, “west coast”), countries (*e.g.* “USA”, “U.S.”, “america”), or abstract concepts meant to be jokes (*e.g.* “none of your business”). Previous work has used the location fields on Bluelight and drugs-forum user profiles to geolocate data to the county- (*52*) or country-level (*53*).

Other platforms contain similar location fields, though these are less frequently populated than on forums. Schwartz *et al.* processed these optional location entries on X to create the County Tweet Lexical Bank, a dataset of tweets geolocated at the county level (*54*). Several groups have leveraged the County Tweet Lexical Bank and datasets assembled by similar methods for opioid-related (*8*, *9*, *26*, *55*, *56*) and non-opioid-related (*57*, *58*) work. Others have used named entity recognition to extract unambiguous place names from social media text (*59*–*62*).

Several groups have used the hashtag search functionality on TikTok to obtain content relevant to geographic regions (*e.g.* specific countries) (*63*, *64*). Similarly, Hu and Conway pulled text from country-specific subreddits as a proxy for geolocation (*65*); Delbruel *et al.* examined the association between YouTube video tags and geolocation (*66*); and Goyer *et al.* used keyword search to identify Reddit and X posts relevant to Canada and manually identified Canadian Facebook groups from which to extract content (*67*).

A key feature of social networks is that users are connected (through “friendship”, “following”, *etc.*) to other users with whom they often share characteristics or interests; sometimes we can infer the location of a user based on the locations of users to whom they are connected. For example, someone who almost exclusively follows people based in New York City on Instagram is likely to also be based in New York City. This line of reasoning has been used extensively for X (*68–70*) because it requires the social network being partially labeled with geolocation.

Examining a social media user’s posts beyond the topic of interest and profile information beyond an explicitly stated location can also yield a proxy for geolocation. This strategy is often used for Reddit as it is facilitated by explicit subforum (subreddit) topics and post titles, key features of this platform. Several groups have used the assumption that if a Reddit user frequently posts in a city-specific subreddit, then they are likely to live or spend a significant amount of time in that city (*e.g.* posting frequently in r/sanfrancisco implies living in San Francisco) (*2*, *71*, *72*). Others have also searched for posts with the topic of “where are you from?” or instances of the phrase “I live in…” (*72*, *73*) and leveraged user “flairs” (tags that Reddit users can add to their username) (*72*) to geolocate Reddit users.

Numerous packages for predicting geolocation from social media data exist. Free packages used previously for geolocating social media text (*7*, *74*) include Carmen (*75*) and geopy (*76*); paid services used previously for geolocating social media text (*77–79*) include Iconosquare (*80*), Brandwatch (formerly Crimson Hexagon) (*81*), and Reputation (formerly Nuvi) (*82*). Additionally, many other groups have created geolocation inference methods (*32*, *83–86*). While these methods can facilitate geotagging, the accuracy of such methods decreases with time (*87*).

A final strategy to obtain geolocation estimates is to link users to accounts on a different platform that does have geolocation. For example, while Tumblr does not provide geolocation, Tumblr users can share their posts to X. Xu *et al.* leveraged this cross-posting to obtain geolocation information for Tumblr users (*62*, *88*).

We summarize geolocation inference strategies for each of the shortlisted platforms in Table 1.

### Prior use in research literature

Of all shortlisted platforms, X and Reddit were the most commonly used in existing literature. The work using these two platforms ranges from correlating opioid-related discussion volume and opioid-related overdose death rates (*2*, *7–10*, *25*, *26*, *55*, *56*), characterizing trends and themes in online discussion of opioids, opioid use, and OUD treatment (*4*, *21–24*, *55*, *59*, *72*, *89–118*), and characterizing public sentiment towards the opioid epidemic (*23*, *67*, *79*, *95*, *109*, *119*, *120*). Many groups have created models to identify posts on X and Reddit related to opioids (42,43,109,134–138), and one group created a pipeline to infer demographic information of the identified cohort (*126*). Others have used these platforms to characterize factors that influence opioid use, recovery, and the opioid epidemic generally (*6*, *8*, *127–131*), with particular interest shown to the impact of the COVID-19 pandemic (*90*, *107*, *132–140*) and co-use between opioids and other drugs (*141*, *142*).

The platforms with the next most volume of prior work were Instagram and Facebook. Previous work used these data to examine aspects of opioid use (*143–145*), to characterize content related to drug sales (*146–149*), to identify emerging psychoactive substances (*108*), and to study public reactions to the opioid epidemic (*67*, *150*, *151*). Though Instagram is an image-focused platform, nearly all studies used text from captions and comments for analysis. Facebook text data was obtained from public groups and pages.

YouTube, Tumblr, and drugs-forum have seen modest usage in text-based research focused on the opioid epidemic. YouTube comments have been analyzed with NLP techniques to characterize sentiments toward opioid use (*152*) and the opioid epidemic (*153*). YouTube and drugs-forum were both used to characterize misinformation related to OUD medications (*92*, *118*). Catalani *et al.* searched YouTube and Tumblr for content about emerging psychoactive substances (*108*). Others have used Tumblr text data to detect opioid sales (*146*, *154*, *155*). Paul *et al.* (*53*) and Lee *et al.* (*156*) used drugs-forum text data to establish correlations with NSDUH survey data. Drugs-forum has also been used to monitor other drugs (*157*, *158*).

We did not find relevant prior work using Pinterest, TikTok, Bluelight, or LinkedIn. Previous work used Pinterest and TikTok to study portrayals of drugs other than opioids (*159*–*166*). Vosburg *et al.* (*167*) and Soussan and Kjellgren (*168*) used Bluelight in the context of recruiting participants for studies related to opioid use. Our PubMed query returned no results related to opioid research for LinkedIn.

## Discussion

Social media data signals correlate with trends in the opioid epidemic. Although existing research has leveraged social media platforms to analyze such phenomena, most research has focused on only a few of the platforms currently in use. We have characterized the utility and accessibility of platforms with potential for monitoring opioid-related discussion to motivate future research and give a more complete perspective of emerging trends in the North American opioid epidemic.

We found that all eleven shortlisted platforms contain notable volumes of opioid-related discussion. Beyond total volume of opioid-related content, other factors affecting utility in an opioid epidemic surveillance system include degree of censorship, user base demographics, and geolocatability. In addition, the accessibility and stability of these data sources affect their utility for public health research or surveillance. While we highlight APIs for shortlisted platforms, not all drug-related discussion or user metadata are available through official APIs. Recently, some platforms have shifted from freely available APIs to paywalled versions, which may be cost-prohibitive for large-scale projects. Evolving user language makes it challenging to capture relevant discussions, as with the emergence of algospeak. While all shortlisted platforms are potential sources for surveillance methods, we highlight TikTok, Facebook, and YouTube as underutilized platforms with significant opioid-related content. Reddit and X have a rich existing body of research and remain valuable data sources, but as their access has become restricted, additional sources are necessary.

In this work, we did not differentiate between opioid mentions in the context of self-disclosure of opioid use and other types of mentions because this distinction is not necessary for correlation with opioid mortality rates or uncovering themes in perception of opioids (*2*). Different downstream use cases will require different decisions on how to narrow the scope of opioid mentions for analysis.

### Platform users, contents and dynamics affect research utility

Platforms that allow open discussion of drug-related topics (*e.g.* Bluelight, drugs-forum) or that grant pseudo-anonymity (*e.g.* X, Tumblr) afford freedom in discussing stigmatized topics. This increases the amount of opioid mentions on a platform, as we observed. Platforms with high levels of moderation may spur algospeak usage. This phenomenon has been described in the literature with respect to TikTok (*169–171*).

Demographics play a key role in selecting social media platforms for surveillance of the opioid epidemic. For example, the user base for TikTok skews to younger age groups. In a Statista survey, 67% of respondents aged 18-19 and 56% of respondents aged 20-29 reported TikTok use, compared to 38% of 40-49 year olds (*172*). Individuals who initiate opioid use at a younger age are more susceptible to substance use disorders (*173*); monitoring discussions around opioid use in younger people could help inform preventative programs. Facebook skews older, with 75% of 30-49 year olds reporting that they use the platforms as opposed to 67% of 18-29 year olds (*174*). Platform demographics also vary by gender. A greater proportion of women than men report using Facebook, Instagram, TikTok, and Pinterest, whereas a greater proportion of men than women report using X and Reddit (*174*). Using multiple social media platforms in order to monitor different demographics may be required in the context of the opioid epidemic. The subset of platform users discussing opioids may have a different demographic makeup than the overall platform user base; accordingly, demographic attributes such as race, age, and gender for opioid-mentioning users are critical for evaluating whether populations disproportionately impacted by the opioid epidemic are represented. Such pipelines have been developed for social media generally (*175–178*) and in the context of substance use (*126*). Many of these tools have demonstrated good performance at the population scale. However, they are not suitable for estimating demographic attributes (especially race and gender) of individual social media accounts, as these are sensitive and may require additional human subject research protections. They may also introduce stereotype, error, and erasure of minority identities.

Most prior work in social media pharmacovigilance has been conducted using X and Reddit. They have high volume of content (both in total and drug-related), formerly freely accessible APIs, and inferrable or explicitly-provided geolocation data. However, other platforms used less in research share these attributes. We have identified potential geolocation inference strategies for all 11 shortlisted platforms. Facebook, YouTube, TikTok, and LinkedIn emerged as platforms with high total volumes of opioid-related discussion but low prior use in the literature. According to Pew Research Center, Facebook and YouTube are the most-used online platforms by Americans as of January 2024 (*174*), and TikTok has emerged as a popular platform for youth. We found that LinkedIn’s high volume of opioid-related discussion was the result of people sharing research and educational resources related to opioids.These four platforms may provide insight supplementary to that from X and Reddit.

### API access to social media data can be limited

It is not always possible to retrieve all data collected by a social media platform through its API. Many social media platforms infer geolocation and demographic information from user activity, but will omit such identifiers from available datasets. Some platforms only allow queries from a subsample of data rather than the full historical archive. Alternatives to APIs for obtaining data include manual search, web scraping of public webpages, or soliciting donation of private data directly from users (*67*, *145*, *150*, *151*, *179*). Web scraping and third-party APIs may violate a platform’s terms of service.

Many platforms allow users to post content that is not publicly viewable and cannot be obtained through APIs or web scraping. Users may discuss opioid use on social media among their private networks, but share more filtered accounts publicly. If there are differences in users that engage in public versus private discourse, data will be subject to selection biases. On platforms with pseudo-anonymous users, private posts may be less prevalent.

### Changing business models affect platform availability and stability

The policies of social media companies are rapidly evolving, leading to unstable data access for research. For example, efforts to monetize the primary Reddit API effectively disabled the third-party Pushshift API, which had previously facilitated the high volume of prior research using the platform.

Data instability poses a challenge to surveilling opioid overdoses longitudinally. Incorporating other data sources could make systems more robust to gaps in data access. Ideally, mechanisms for working directly with social media companies would establish more stable data access.

### Limitations

The internet is dynamic. The way in which people use social media platforms to talk about opioids will change with time, and this must be accounted for when analyzing this type of data. Informal language constantly evolves, and the terms used here may not be in use in the future. Here, we detailed a procedure that used generative models to create a list of terms; we believe that these methods could be used in the future to update informal opioid term lists. However, workflows based on generative models cannot capture emergent slang terms that differ from those in their training data. To circumvent this limitation of a generative AI approach, the best strategy may be to identify drug-related terms directly from the dataset.

The use of social media data requires careful ethical consideration – particularly with respect to privacy. Even publicly-visible social media posts related to opioids are sensitive in nature and may not be suitable to share via research publication because they relate to a vulnerable population. Social media data mining may also disrupt user trust in social media and may lead to altered behavior. Although there is no standard ethical guidance for research on mined social media data (*180*), good practices include redacting usernames or other identifiable information from shared data, paraphrasing posts instead of publishing direct quotes, and actively engaging with community moderators. Research should not contribute to stigmatization, individual-level surveillance, and harm to vulnerable populations. The ethical issues inherent to analysis of social media data, and suggestions on how to conduct such research ethically, are addressed in (*181*).

We used Google search queries as an estimate of the volume of public opioid-related discussion. We use this proxy rather than accessing and processing text-based content from each platform, which would require substantial computing resources. These estimates may be influenced by how Google indexes web pages. Platforms owned by Google, such as Youtube, may be overrepresented in search results. Other platforms may restrict access to unregistered users, impacting how Google can process their webpages (*182*). We have provided an initial estimate of the volume of opioid-related discussion on each site, but the actual amount of accessible data may vary.

A key limitation of our analysis is its explicit focus on English text data on platforms primarily serving North America. Our choice to focus on North America was driven by the severity of the opioid epidemic in the United States and Canada. However, OUD is a global problem and analysis beyond North America is needed. We also recognize that there are likely substantial opioid-related discussions in languages other than English within North America. According to the U.S. Census Bureau, over 20% of Americans speak a language other than English at home (*183*). Nevertheless, our methods can be repurposed for other countries and languages. For example, to conduct a similar analysis for South America, one should shortlist platforms widely used in South America with significant Spanish and Portuguese language content, as well as construct Spanish and Portuguese term lists with formal and informal terms for the most common opioids in South America. Future research in leveraging social media platforms for pharmacovigilance of the opioid epidemic in regions other than North America and languages other than English would enable a more complete assessment of this worldwide crisis.

Finally, we recognize that social media cannot fully capture all phenomena relevant to the opioid epidemic. Integrating social media data with other sources, such as emergency medical services, drug seizure records, and wastewater analysis, may supplement shortcomings and increase the efficacy of an early-warning system.

## Materials and Methods

### Experimental Design

The objective of this study was to identify social media platforms that have been underutilized in text-based research monitoring the opioid epidemic but contain opioid-related discussion and have characteristics favorable for large-scale research. The study consisted of three components: the broad platform identification stage, the platform shortlisting stage, and the platform characterization stage. The prespecified criteria and methods are detailed for each stage in their respective subsections below.

### Identifying social media platforms

We compiled a list of social media platforms that may contain content related to opioid use, substance use disorders, and/or addiction treatment and recovery. We used three criteria to select platforms: a platform must either be 1) a general-purpose social platform with widespread use; 2) used previously in research for drug-related surveillance; or 3) a platform with a history of drug-related transactions.

With the first criterion, we sought to include all current major social media platforms, as any large general-purpose platform for online discourse is likely to include mentions of opioids. We included all platforms with more than 100M monthly active users worldwide, as per the list compiled by Wikipedia (*184*). We also included all platforms that the Stanford Digital Economy Lab identified as “digital goods” due to their widespread use and relevance (*14*).

We anticipated that smaller platforms with focus on drug-related conversation could also be useful data sources. For the second criterion, we searched relevant literature for studies using social media as a data source for drug-related surveillance work, “social media”, and “pharmacovigilance”. We identified studies and reviews using social media and discussion forums for surveillance of illicit drug use (*146*, *186–188*) and of general adverse drug reactions (*53*, *186*, *189–196*). We included all platforms used by the identified studies and reviews.

For the third criterion, we sought to include platforms with a history of illicit drug transactions. We identified popular online marketplaces and social messaging platforms known to have previously been used to coordinate illicit drug sales (*89*, *197–203*)

### Creating platform shortlist

We determined if each platform 1) had an active web domain or mobile application, 2) met the Knight First Amendment Institute definition of social media (*204*), 3) had a primary function other than private messaging, 4) was based in the US/Canada or had English as the default language for US-based users, and 5) returned more than 25,000 Google search hits for a set of opioid-related terms. Platforms that met all five criteria were shortlisted for evaluation. A description of the shortlisting process can be found in **Supplementary Text S1**.

### Measuring the volume of opioid-related discussion

We used the number of hits returned by Google search results when querying opioid-related terms to approximate the amount of opioid-related discussion on each platform. While an imprecise approach, Google search matches allowed us to characterize the general volume of opioid mentions on each platform and make comparisons between platforms in an efficient, standardized manner.

We formatted the queries so as to only yield results from the specific platform’s domain, *e.g.* when assessing Facebook, we limited results to only be those from facebook.com. We assembled three lists of opioid-related terms: “formal,” “informal,” and “algospeak.”

The “formal” opioid term list includes generic names and brand names of opioids that are currently key drivers of the opioid epidemic in North America (prescription opioids and fentanyl). While heroin drove the second wave of the North American opioid epidemic, and remains a primary cause of opioid overdose deaths in other regions, we excluded it from our term lists in an effort to narrow our focus to the largest threats in the current North American opioid epidemic. These terms are the same as those used to select for platforms with high opioid discussion in the shortlisting phase (**Supplementary Text S1**, **Supplementary Text S2**).

The “informal” opioid term list includes slang, misspellings, and other brand names. Because language on social media is often informal, such terms are important for capturing the full scale of opioid-related discussion. We previously found that GPT-3 (*205*) is able to generate slang for drugs of addiction at scale (*206*). We took validated GPT-3–generated slang terms for five opioids prominent in the current North American opioid epidemic (four prescription opioids: codeine, morphine, oxycodone, and oxymorphone; and fentanyl) and removed terms that had common non-drug meanings. To keep the scale similar to the formal term analysis, we reduced the list to the 20 terms most frequently generated by GPT-3 (**Supplementary Text S3**).

“Algospeak” is a phenomenon in which users of social media platforms purposefully alter words to evade banning and censorship (*169*). For example, a poster on social media may refer to fentanyl as “f3ntanyl.” The set of terms generated by GPT-3 did not include algospeak. We created an “algospeak” term list using GPT-4 (**Supplementary Text S4**, **S5**).

We used the Google Search API with default parameters to query for the number of English-language hits specific to a given social media website for each of the formal opioid terms, informal opioid terms, and algospeak opioid terms. To reduce variation, all search queries were executed on January 29, 2024. We tabulated the total number of hits per list to estimate the total volume of opioid discussion on each of the shortlisted platforms. Additionally, we normalized the number of opioid-related hits by the number of hits returned for queries of “household terms”, chosen from Corpus Of Contemporary American English to represent the most common nouns used in the English language (**Supplementary Text S6, S9-11**). This ratio represents how prominent opioid discussion is relative to other content on the platform, allowing for comparison between platforms with varying popularity. However, we note that because the success of downstream analysis depends upon having sufficient opioid-related text data, popular platforms with a greater total number of opioid-related hits may be preferable to smaller platforms enriched for opioid-related discussion. In the following analyses, we combine the informal and algospeak normalized ratios, as algospeak is a special case of informal language.

### Content restrictions and censorship policies

We analyzed the content restriction and censorship policies of social media sites by referencing the sites’ terms of use and user agreements.

### Evaluating data accessibility for academic research purposes

We sourced information regarding data access for research purposes from public information on each website. We used this information to determine whether geolocation is provided for any accessible data.

### Assessing prior use of platforms in research literature

We conducted PubMed searches to identify literature relevant to each of the shortlisted platforms that focuses on applications to the opioid epidemic. We searched for articles with the name of the social media platform in the title or abstract (for less commonly studied social media platforms, this criterion was broadened to presence anywhere in the article) and either “opioids”, “opioid”, “opiates”, or “opiate” in the title or abstract.

Example: (twitter[Title/Abstract]) AND (opioids[Title/Abstract] OR opioid[Title/Abstract] OR opiates[Title/Abstract] OR opiate[Title/Abstract])

In addition, we substituted the term “social media” in place of the platform name. Search queries were performed in March 2024; our results cover papers published until that time.

We defined relevant literature as papers that describe a primary research study analyzing data originating from at least one of our 11 shortlisted platforms to obtain information related to opioid use. We excluded review papers, studies describing social media-based interventions, and studies that only used social media as a participant recruitment tool. We evaluated the abstract and, if required, the full text of all papers returned by the described PubMed queries to determine if they met these criteria.

### Statistical Analysis

Not applicable.

## Data Availability

All data and code produced are available online at https://github.com/kristycarp/opioid-social-media-platforms.

## Acknowledgments

The authors would like to thank Salvatore Giorgi, Shashanka Subrahmanya, Aadesh Salecha, and Rhana Hashemi for insightful conversations and assistance.

## Funding

National Institutes of Health grant R21DA057598

Microsoft Accelerating Foundation Models Research Initiative

National Science Foundation grant DGE-1656518 (KAC)

National Institutes of Health grant T32HL151323 (ATN)

Stanford Biochemistry Department (DAS)

National Science Foundation grant 2019286895 (DAS)

Stanford Summer First Fellowship (IAS)

Sarafan ChEM-H CBI Program Award (IAS)

Department of Veterans Affairs Health System Research Service Award RCS 04-141-3 (KH)

Stanford Institute for Human-Centered AI (JCE, RBA, AL)

Chan Zuckerberg Biohub (RBA)

## Author contributions

Conceptualization: RBA, JCE

Methodology: KAC, ATN, DAS, JCE, RBA

Investigation: KAC, ATN, DAS, IAS

Visualization: KAC, ATN, IAS, JCE

Supervision: JCE, RBA, KH, AL, MVK

Writing—original draft: KAC, ATN, DAS

Writing—review & editing: KAC, ATN, IAS, KH, AL, MVK, JCE, RBA

## Data and materials availability

Code for the analyses contained here is available at https://github.com/kristycarp/opioid-social-media-platforms. All other data are present in the main text and the Supplementary Materials.

