## Supplementary Text for "Which social media platforms facilitate monitoring the opioid crisis?"

Kristy A. Carpenter *et al.*

**This PDF file includes:**

Supplementary Text S1 to S11

Fig. S1

Tables S1 to S2

**Other Supplementary Materials for this manuscript include the following:**

Data S1 to S2

Supplementary Text

S1. Details of platform shortlisting methods

We considered a platform to be active if we were able to access a website on an internet browser or if a mobile application was available for download. We chose to restrict our platform list to include only those that have an active website or mobile application to identify opportunities for future prospective research.

We classified platforms as social media following a definition used by the Knight First Amendment Institute at Columbia University (*204*): social media is defined as “a digital space that combines communicating or sharing media with aspects of social networking sites,” and only includes platforms that are primarily social in nature (e.g. excluding news sites with comment sections, marketplaces, and content subscription services).

We considered a platform to be primarily for private messaging if its only features were person-to-person or group messages. We excluded these platforms as their data is not accessible for research use.

We only included platforms that were based in the United States or Canada, or that used English as the default language for a user based in the United States. While opioid misuse and addiction is a global crisis, we chose to focus on North America given the severity of the opioid epidemic in the region (*14*, *15*).

To estimate how much opioid-related discussion exists on each platform, we used the Google Search API to query for a set of opioid-related keywords (**Supplementary Text S3**). We selected only keywords that are specific to opioid use. If the query returned more than 25,000 hits for a platform, we considered that platform to have a high volume of opioid-related discussion.

Two authors (ATN, IAS) independently assessed each platform. The authors compared their assessments, identified discrepancies in their judgements, and independently completed a second review of platforms for which there were disagreements. The authors then discussed their secondary evaluations until they agreed on the assessment of each platform.

S2. Formal opioid term list

fentanyl, opioids, opiates, morphine, codeine, oxycodone, oxymorphone, mscontin, percocet (n = 9)

S3. Informal opioid term list

sublimaze, duragesic, fentanil, sufentanil, fentanylum, fentora, thebaine, codiene, roxanol, kadian, oxycontin, roxicodone, roxicet, endocet, endocodone, oxyir, oxynorm, hydrocodone, vicodin (n=19)

S4. Algospeak opioid term list

paink!ller, f3nt@nol, cod3in3, c0d0n3, f3nt4nol, p@1nk!ller, oxy80s, 0xy80, m0rph!n3, m3rph0n3, m0rf3n, c0d3in, 0xyc, s!zzurp, 0pana, 0xym0rph0ne, num0rph@n, m0rf33n (n = 18)

S5. Algospeak generation protocol

We used the OpenAI Chat Completions API from Microsoft Azure to generate our algospeak slang terms. We used the gpt-4 model and the “2023-05-15” Azure API version. We set the temperature to 0.7.

We gave the model the following system message as context:

“You are an AI assistant that helps people find information. You are particularly hip with online slang and know everything about how people talk on social media platforms like Facebook, Twitter, Reddit, and TikTok.”

We used the following prompt template, which was developed through iterative prompt engineering in a sandbox environment:

“We are playing a game where I give you a word, and then you give me algospeak terms for that word. The algospeak must be in the format such that none of the included words would be present in an English dictionary. I will give you an example; please continue each of these comma-separated lists:

1. dog: d0g, pupper, doggo

2. die: d!e, unalive, d!3

3. [INSERT QUERY DRUG EXAMPLE]

4. friend: fr13nd, @mig0, frend

In your response, only give the four lists (for dog, die, [INSERT QUERY DRUG], and friend) with comma-separated terms. Do not include any other text.”

Using this template, we created prompts for each of the five opioids for which to generate algospeak terms. We replaced [INSERT QUERY DRUG] with one of codeine, fentanyl, morphine, oxycodone, or oxymorphone. We replaced [INSERT QUERY DRUG EXAMPLE] with the corresponding algospeak example list for the selected drug:

codeine: c0dein3, c0dine, cod eine

fentanyl: f3ntanyl, f3nt, f3ntanil

morphine: m0rphine, morf1ne, m0rph1n3

oxycodone: 0xy, 0xyc0d0n, p3rcs

oxymorphone: 0xym0rph0ne, 0pana, num0rph@n

All example algospeak terms were human-generated based on observed internet language patterns.

We acknowledge that this prompt setup, including the system context statement, could potentially find improvement with modification, particularly ensuring that all language reflects current common internet text patterns. However, because prompt engineering is a process that can iterate interminably if one is searching for an optimal prompt, and because we saw that this prompt produced desirable results, we have elected to use this prompt for a production run without undertaking additional optimization experiments.

We submitted each drug query to the Chat Completions API 100 times and processed the results to extract all generated algospeak drug terms. This yielded 300 codeine terms (74 unique), 303 fentanyl terms (167 unique), 310 morphine terms (169 unique), 310 oxycodone terms (143 unique), and 356 oxymorphone terms (198 unique). We eliminated any terms only generated once in an attempt to remove noise. This left 26 codeine terms, 32 fentanyl terms, 35 morphine terms, 47 oxycodone terms, and 41 oxymorphone terms. We also eliminated any terms that did not include numbers or special characters, as those without those characters did not fit our definition of “algospeak.” We eliminated any terms that had a corresponding “non-algospeak” term that did not have an opioid-specific meaning (*e.g.* the non-algospeak equivalent of “f3ntanil” is “fentanil”). To reduce the scale of the list to a similar scale as that of the formal and informal opioid terms, we kept only terms generated more than four times, which yielded a final list of 18 algospeak terms.

S6. Common noun / “household term” list

time, way, life, place, friend, idea, name, death, son (n=9)

S7. Current content restriction policies, by platform (as of February 1, 2024)

*Bluelight*

“Bluelight neither condemns nor condones the use of drugs. Rather, we accept that drug use will always exist irrespective of legal status or societal norms. While there is no truly safe way to use drugs, we understand that prohibition and abstinence are not realistic or desirable solutions for everyone, nor have they been adequate in addressing the serious public health concerns associated with drug use… These forums invite visitors to discuss addition and sobriety in a non-judgmental setting, share recovery resources and encourage members to seek help." (<https://bluelight.org/xf/pages/BL_AboutUs/>)

“[Y]ou may not use Bluelight in any way, shape or form for unlawful purposes, including, without limitation: attempting to solicit, obtain sell or supply contraband substances or substances of a quasi-legal status or requesting information on how to do so…” (<https://bluelight.org/xf/pages/BLUA/>)

*Drugs-forum*

“Requesting or offering illegal substances is not allowed. This includes chemicals on the UN red list, DEA List I or scheduled precursors as well as thresholds or above quantities of DEA List II chemicals. This will get you banned instantly… Requesting, offering or trading sources for illegal substances is not allowed. This will get you banned instantly… Do not advertise any commercial websites or products." (<https://drugs-forum.com/threads/the-rules.298373/>)

“As a result of our strict moderation, Drugs-Forum offers a safe space for discussion of all aspects of recovery and drug use, both medical and recreational.” (<https://drugs-forum.com/>)

*Facebook*

“Do not post content that: Attempts to buy, sell, trade, co-ordinate the trade of, donate, gift or ask for high-risk drugs. Admits to buying, trading or co-ordinating the trade of high-risk drugs by the poster of the content by themselves or through others. Admits to personal use without acknowledgment of or reference to recovery, treatment, or other assistance to combat usage. This content may not speak positively about, encourage use of, coordinate or provide instructions to make or use high-risk drugs. Coordinates or promotes (by which we mean speaks positively about, encourages the use of, or provides instructions to use or make) high-risk drugs. Do not post content that: Attempts to buy, sell, trade, co-ordinate the trade of, donate, gift or asks for non-medical drugs. Admits to buying, trading or co-ordinating the trade of non-medical drugs by the poster of the content by themselves or through others. Admits to personal use without acknowledgment of or reference to recovery, treatment, or other assistance to combat usage. This content may not speak positively about, encourage use of, coordinate or provide instructions to make or use non-medical drugs. Coordinates or promotes (by which we mean speaks positively about, encourages the use of, or provides instructions to use or make) non-medical drugs.”  (<https://transparency.fb.com/policies/community-standards/restricted-goods-services/>)

*Instagram*

“[B]uying or selling non-medical or pharmaceutical drugs are also not allowed. We also remove content that attempts to trade, co-ordinate the trade of, donate, gift, or ask for non-medical drugs, as well as content that either admits to personal use (unless in the recovery context) or coordinates or promotes the use of non-medical drugs.” (<https://help.instagram.com/477434105621119>)

*LinkedIn*

“Do not promote, sell or attempt to purchase illegal or dangerous goods or services. We don’t allow content that facilitates the purchase of illegal or dangerous goods and/or services… We also don’t allow content depicting or promoting instructional weapon making, drug abuse, and threats of theft." (<https://www.linkedin.com/legal/professional-community-policies>)

*Pinterest*

“Pinterest isn’t a place for trading or selling of certain regulated goods—products or substances that can cause harm when used, altered or manufactured irresponsibly—or for the display or encouragement of dangerous activities. We limit the distribution of or remove such content and accounts, including: Individuals and unlicensed retailers offering to sell, purchase or trade alcohol, tobacco, drugs and weapons, including firearms and accessories, firearm parts or attachments, or ammunition; Content from or about unverified, unapproved or rogue online pharmacies; Offers, attempts or instructions to bypass purchasing laws and regulation; Instructions for creating lethal or toxic substances…" (<https://policy.pinterest.com/en/community-guidelines>)

*Reddit*

“Keep it legal, and avoid posting illegal content or soliciting or facilitating illegal or prohibited transactions." (<https://www.redditinc.com/policies/content-policy>)

“Community content tags are tags that moderators add to their communities to let redditors know what kind of mature content is in that community… Reddit moderators set the content tags for their communities by taking a quick survey about how a community’s posts and discussions currently involve the following mature themes: Amateur advice; Alcohol & tobacco; Drug use; Gambling; Guns & weapons; Nudity; Profanity; Sex & eroticism; Violence” (<https://support.reddithelp.com/hc/en-us/articles/360048185132-What-are-community-content-tags-and-how-do-they-work>)

*TikTok*

“While adults make personal choices about how they engage with alcohol, drugs, and tobacco, we recognize that there are risks connected to trading and using these substances. We do not allow showing or promoting recreational drug use, or the trade of alcohol, tobacco products, and drugs. We also recognize that using these substances can put young people at a heightened risk of harm. We do not allow showing or promoting young people possessing or consuming alcohol, tobacco products, and drugs. Content is age-restricted and ineligible for the For You Feed (FYF) if it shows adults consuming excessive amounts of alcohol or tobacco products.”

“NOT allowed: Showing or promoting young people possessing or consuming alcohol, tobacco products, drugs, or other regulated substances; Showing or promoting adults consuming drugs or other regulated substances for a recreational purpose; Showing the misuse of common household items or over-the-counter products to get intoxicated, such as antihistamines, nutmeg, nitrous oxide canisters, and sniffing glue; Providing instructions on how to make homemade spirits, drugs, or other regulated substances; Facilitating the trade or purchase of alcohol, tobacco products, drugs, or other regulated substances.”

“Allowed: Raising awareness about substance misuse and sharing recovery stories; Advocating for the reform of drug policies and regulations.”

(<https://www.tiktok.com/community-guidelines/en/regulated-commercial-activities>)

*Tumblr*

“Don’t use Tumblr for any kind of exchange of regulated drugs, substances, devices, goods, or weapons. Don't use Tumblr to buy them, sell them, trade them, or to share instructions for manufacturing them." (<https://www.tumblr.com/policy/en/community>)

*X (Twitter)*

“You may not use our service for any unlawful purpose or in furtherance of illegal activities. This includes selling, buying, or facilitating transactions in illegal goods or services, as well as certain types of regulated goods or services." (<https://help.twitter.com/en/rules-and-policies/regulated-goods-services>)

*YouTube*

“[T]he following content isn’t allowed on YouTube: Hard drug use or creation: Hard drug use or creation, selling or facilitating the sale of hard or soft drugs, facilitating the sale of regulated pharmaceuticals without a prescription, or showing how to use steroids in non-educational content. Generally, hard drugs are defined as drugs that can lead to physical addiction, such as cocaine or opioids. Soft drugs include marijuana and salvia.”

“Here are some examples of content that’s not allowed on YouTube… Content instructing how to purchase drugs on the dark web… Including a link to an online pharmacy that does not require prescriptions. Content that promotes a product that contains drugs, nicotine, or a controlled substance. Displays of hard drug use: Non-educational content that shows the injection of intravenous drugs like heroin or huffing/sniffing glue. Making hard drugs: Non-educational content that explains how to make drugs.” (<https://support.google.com/youtube/answer/9229611>)

S8. Current data access policies, by platform (as of December 1, 2023)

*Bluelight*

Bluelight.org has a dedicated research portal in alignment with its mission to be a resource for research into drug use and harm reduction. The research portal offers services to researchers and is a starting point for collaboration with the platform. The organization notes that the discussions posted on their site are “cited in the scientific literature published on novel and emerging drugs." Discussions are public and available for research use. However, user demographic data is released in a Bluelight census report, with the most recent being in 2018.

Bluelight contains strict guidelines about data usage:

"If you would like to use Bluelight content as data in your research project, we respectfully wish to remind you that unauthorized reproduction or use of content from Bluelight is explicitly prohibited by our User Agreement. Projects of this nature can be conducted with our permission and ideally in partnership with us. We welcome new ideas about research that uses our data and we encourage you to contact our research team to discuss your ideas further."

"All messages posted become the exclusive property of Bluelight.org. Unauthorized reproduction or use is prohibited. Bluelight partners with both public and private research organizations and we encourage researchers to contact us for more information on our research standards and protocols."

*Drugs-forum*

Drugs-forum.com has no available API. The content was generally available for research purposes, as it was previously archived on Substance Abuse and Mental Health Data Archive.

*Facebook*

Facebook currently operates under an open research & transparency (FORT) policy, which provides curated research datasets (*e.g.* ad targeting transparency, URL sharing, civic engagement data) through a research-specific API. There is an application process to access these data, and access is structured through a remote computing instance where researchers can access and analyze through Jupyterlab. Facebook claims to provide "near real-time data as well as billions of historical data points including raw, anonymized data from public forums on Facebook including Groups, Events & Pages."

*Instagram*

Instagram offers an API for app developers. Instagram shares basic user demographics including self-reported gender, age and broad location with developers, although access for research purposes varies. There are several third party API products for accessing these data, although Instagram warns that "there are a handful of pre-built web scraping packages for Python and R that are specifically designed to collect data from Instagram. Usage of these tools will violate the Terms of Service on the account you use for the scraping and it may be banned from the platform."

*LinkedIn*

LinkedIn offers an API, with different versions for free and premium users. Data includes post content and metadata, user information, learning platform data, and job listings. LinkedIn claims that the API itself allows third parties to retrieve information from LinkedIn without any restrictions.

*Pinterest*

Pinterest has no academic or research API, but does offer an analytics interface. Information shared includes metadata regarding pins, including data abouts apps, products, recipes, and articles tied to each pin. User related information is collected, but is used for internal purposes, with only public information such as username being available in the API. There are several third-party APIs that claim to have access to demographic data such as user age and location, but these are unverified and may violate the Pinterest user agreement.

*Reddit*

Reddit data is limited in the demographic outcomes that it can provide. Reddit provides posts, comments, and user information such as handle name and number of posts. Data was formerly available through Pushshift, a third-party API. However, since Reddit started charging for API use, Pushshift has gone out of operation. Reddit has its own API, but it is rate limited to 100 queries per minute.

*TikTok*

TikTok has a research API available to people who have "demonstrable academic experience and expertise in the research area specified in the application," are employed at a non-profit academic institution, and have a clearly defined research proposal. PhD students need to submit an endorsement letter from an advisor. TikTok shares "all the videos that are: made public by a creator who is aged 18 and over; are posted in the regions of US, Europe and Rest of the World; and do not belong to Canada."

*Tumblr*

Tumblr provides a first-party API with no dedicated research or academic product. Current data from the API includes posts, comments, hashtags as well as usernames and users post data such as volume and number of posts.

*X*

X provides a first-party API, with products that vary in cost and data availability. There is an Academic Research product available to users that are affiliated with an academic institution and have a clearly defined research objective. Data available through both APIs are rapidly changing in the current landscape. Currently, the Academic Research API provides “free access to the full history of public conversation via the full-archive search endpoint, which was previously limited to paid premium or enterprise customers.”

*YouTube*

YouTube has an open-access first-party API as well as a scalable Research API. The research API allows for greater volume of data access but requires an application procedure. It does not allow access to private data but does grant “expanded access to global video metadata across the entire public YouTube.” The data that is available is largely centered around videos, including content, views, and subscriptions to channels. Youtube also offers an audience demographic option that allows content creators to see the age, gender and geography through their analytics portal, although access to these data is limited.

S9. Equation 1: Per-platform Formal Normalized Ratio

$$\text{Formal Normalized Ratio}\left( \text{platform} \right)= \frac{\sum_{\text{formal opioid terms}} \# \text{term hits on platform}}{\sum_{\text{household terms}} \text{\# term hits on platform}}*100,000$$

S10. Equation 2: Per-platform Informal Normalized Ratio

$$\text{Informal Normalized Ratio}\left( \text{platform} \right)= \frac{\sum_{\text{informal opioid terms}} \# \text{term hits on platform}}{\sum_{\text{household terms}} \text{\# term hits on platform}}*100,000$$

S11. Equation 3: Per-platform Algospeak Normalized Ratio

$$\text{Algospeak Normalized Ratio}\left( \text{platform} \right)= \frac{\sum_{\text{algospeak opioid terms}} \# \text{term hits on platform}}{\sum_{\text{household terms}} \text{\# term hits on platform}}*100,000$$

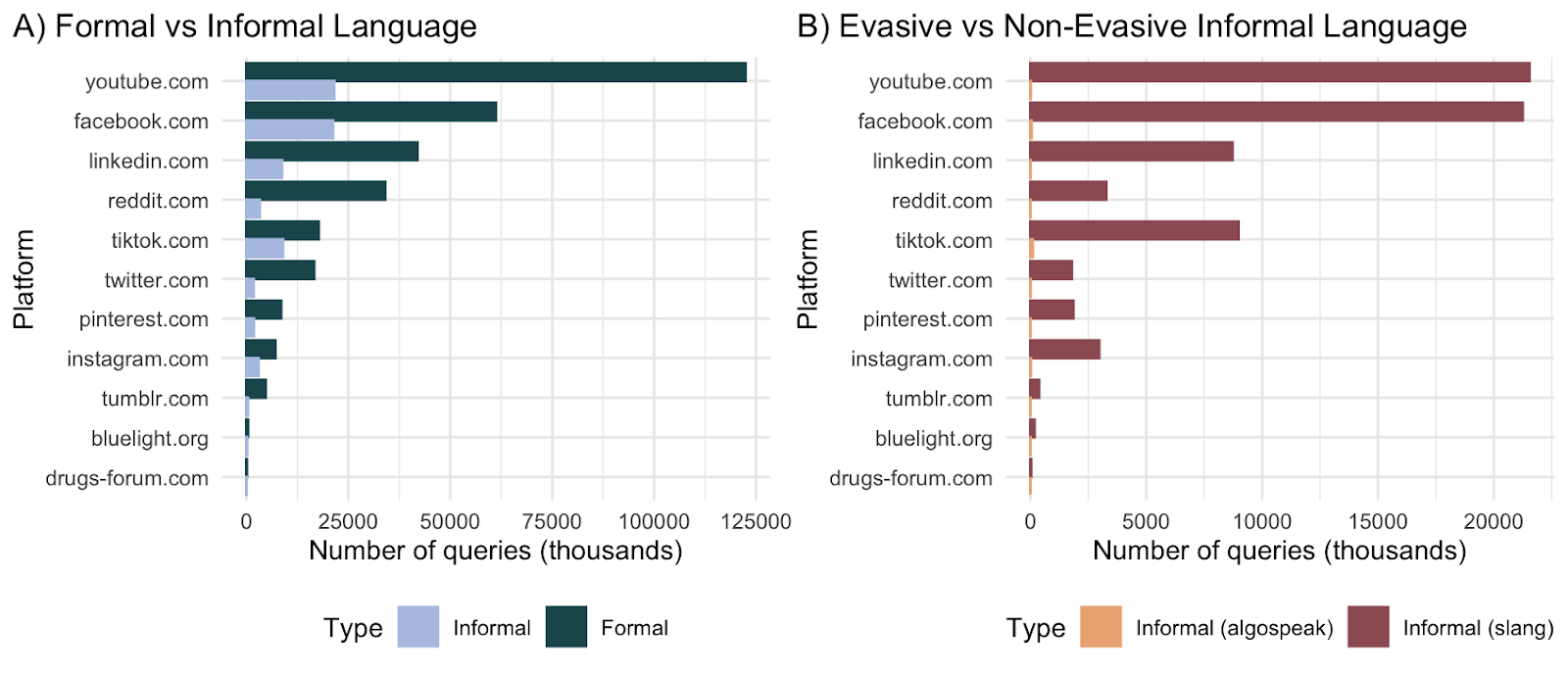

Fig. S1.

Number of query results for collections of opioid-related terms, by social media platform

**(A)** Comparison between formal and informal/algospeak language.

(**B)** Comparison between informal and algospeak language

| Platforms | No. of Household Words Hit (Noun) | Formal Keyword Hits | Informal Keyword Hits | Algospeak Keyword Hits | Informal + Algospeak Keyword Hits |
| --- | --- | --- | --- | --- | --- |
| facebook.com | 1488600000 | 61221824 | 21249847 | 53368 | 21303215 |
| youtube.com | 11322000000 | 122434150 | 21540645 | 14515 | 21555160 |
| instagram.com | 2033900000 | 7096779 | 2971643 | 18962 | 2990605 |
| tiktok.com | 881500000 | 17748308 | 8977614 | 103274 | 9080888 |
| twitter.com | 1105100000 | 16638571 | 1788317 | 9019 | 1797336 |
| pinterest.com | 1183900000 | 8523098 | 1855375 | 3416 | 1858791 |
| linkedin.com | 4816200000 | 41970860 | 8725287 | 6919 | 8732206 |
| reddit.com | 1508000000 | 34062890 | 3272436 | 2034 | 3274470 |
| drugs-forum.com | 109040 | 115063 | 35305 | 0 | 35305 |
| tumblr.com | 325800000 | 4750446 | 376823 | 523 | 377346 |
| bluelight.org | 390800 | 405750 | 180691 | 143 | 180834 |

Table S1.

Raw hit counts from Google Search API across the 11 shortlisted social media platforms. All search queries conducted on January 29 and 30, 2024.

| Platforms | Formal Ratio Scaled | Informal Ratio Scaled | Algospeak Ratio Scaled | Informal+Algospeak Ratio Scaled |
| --- | --- | --- | --- | --- |
| facebook.com | 41127.11541 | 1427.505509 | 3.585113529 | 1431.090622 |
| youtube.com | 10813.82706 | 190.2547695 | 0.1282017311 | 190.3829712 |
| instagram.com | 3489.246767 | 146.1056591 | 0.9322975564 | 147.0379566 |
| tiktok.com | 20134.21214 | 1018.447419 | 11.71571185 | 1030.163131 |
| twitter.com | 15056.16777 | 161.8239978 | 0.8161252375 | 162.6401231 |
| pinterest.com | 7199.170538 | 156.7172058 | 0.2885378833 | 157.0057437 |
| linkedin.com | 8714.51767 | 181.1653793 | 0.1436609775 | 181.3090403 |
| reddit.com | 22588.12334 | 217.0050398 | 0.1348806366 | 217.1399204 |
| drugs-forum.com | 1055236.61 | 32378.02641 | 0 | 32378.02641 |
| tumblr.com | 14580.86556 | 115.6608349 | 0.1605279312 | 115.8213628 |
| bluelight.org | 1038254.862 | 46236.18219 | 36.59160696 | 46272.7738 |

Table S2.

Formal and Informal Normalized Ratios for each of the 11 shortlisted social media platforms, with scaling factor of 100,000. All search queries conducted on January 29 and 30, 2024.

Data S1. (separate file)

Superset of all platforms initially included in analysis, in alphabetical order, with inclusion criteria for each.

Data S2. (separate file)

Evaluation of full platform list.
